# *Plasmodium vivax* relapse, reinfection and recrudescence estimation using genetic data

**DOI:** 10.1101/2022.11.23.22282669

**Authors:** Aimee R Taylor, Yong See Foo, Michael T White

## Abstract

Malaria is a major public health concern. Among the two most important causes of human malaria, *Plasmodium vivax* is the hardest to eliminate, owing largely to its ability to relapse (cause recurrent blood-stage infection following the activation of dormant liver-stage parasites called hypnozoites). Recurrent vivax malaria can also follow a failure to treat a preceding blood-stage infection (recrudescence) and, in endemic settings, a new infectious mosquito bite (reinfection). Understanding the cause of recurrent vivax malaria is critical for disease control efforts, e.g., to estimate the efficacy of a hypnozoiticidal drug, relapse needs to be separated from reinfection and recrudescence. In this report we describe the Pv3R statistical model designed to estimate using *P. vivax* genetic data the probability that a recurrent *P. vivax* blood-stage infection is a relapse, reinfection or recrudescence (the *Pv* 3Rs).

## 1 Introduction

Of the two most important causes of human malaria, *Plasmodium vivax* is the most difficult to eliminate, largely because of its ability to relapse: cause recurrent blood-stage infection via activation of dormant liver-stage parasites called hypnozoites (Olliaro et al., 2016). *P. vivax* recurrence can also be caused by a new infectious mosquito bite (reinfection) and/or failure to treat a blood-stage infection (recrudescence). Knowing the cause of *P. vivax* recurrence is critical for malaria control (e.g., reinfection must be separated from relapse to estimate the efficacy of a hypnozoiticidal drug in an endemic setting). However, at present, there is no manifest way to diagnose the cause, which remains hidden.

*P. vivax* genetic data help elucidate the cause of a *P. vivax* recurrence. Typically, simple rules applied to descriptive analyses of homology, or, more elaborately, of relatedness are used to classify the cause categorically. For example, if alleles detected in a recurrence differ to those detected previously (i.e., they are not homologous), the recurrence is classified either as a reinfection or relapse. Unfortunately, simple homology breaks down when markers are incongruous (some alleles differ and others not). This is where rules applied to more elaborate estimates of relatedness are useful: if parasite genotypes in a recurrence are different but highly related to those in an initial infection, relapse is probably more likely than reinfection.

Regrettably, rules-based classification cannot provide a quantitative measure of the probability of a cause (only a yes/no result) or its uncertainty, which varies with data informativeness and thus data type. In response, a statistical model to infer the probabilities of relapse, reinfection and recrudescence (the 3Rs) using *P. vivax* genetic data was developed (Taylor et al., 2019), demonstrating the feasibility of *P. vivax* 3R genetic inference. In this report we describe an improved version of that model, removing a small amount of bias associated with a simplifying assumption made previously. The improved model, which we hitherto call the Pv3R model, is implemented in the nascent Pv3R R package, which is under development. In the not-so-distant future, we aim to make the Pv3R R package scale to more complex scenarios and further improve the underlying model. However, scalability and additional improvements are beyond the scope of this short report.

On occasion, genetic data alone cannot resolve the 3Rs because unrelated recurrent parasites are compatible with both relapse and reinfection, while clonal recurrent parasites are compatible with both relapse and recrudescence. As such, it is important to allow for other data sources, e.g., time-to-event data, which can be very informative (White, 2011; Battle et al., 2014). This is done by retaining the Bayesian structure of the preceding model, under which prior probabilities were based on posterior predictive draws from a time-to-event model (Taylor et al., 2019). The focus of this report is entirely genetic, however.

## 2 The real world

| Working definitions |  |
| --- | --- |
| **Relapse** | Blood-stage infection following the activation of a hypnozoite derived from a historic inoculation. |
| **Reinfection** | Blood-stage infection derived from a new inoculation. |
| **Recrudescence** | Blood-stage infection following treatment failure of preceding blood-stage infection. |
| **Brood** | Collection of parasites produced when one or more oocysts hatch collectively in a mosquito. |
| **Clones** | Parasites from the same brood that share the same genotype and are thus 100% related. |
| **Siblings** | Parasites from the same brood that have different genotypes but share one or two parental genotypes. Sibling parasites can either come from the same zygote, in which case they necessarily share both parental genotypes (meiotic siblings); or from different zygotes, in which case they can either share both or only one of two parental genotype (regular full and half siblings, respectively). |
| **Strangers** | Parasites from the same brood that do not have a common parental genotype OR parasites from different broods. Parasites from different broods are related insofar as the population across infected humans is inbred and structured. Stranger parasites from the same brood are liable to be more related if their parental parasites were obtained from an infection generated exclusively or in part by mosquito-to-human cotransmission. |
| **Idealised population** | Infinitely large, outbred and panmictic population in which strangers are unrelated and siblings are either $\frac{1}{4}$ or $\frac{1}{3}$, $\frac{1}{2}$ related, depending on whether they are regular half siblings, meiotic siblings or regular full siblings, respectively, thereby implying their parental parasites are strangers. |

### Sexual recombination

Soon after being ingested, pairs of *P. vivax* parasites recombine sexually in the mosquito part of their lifecycle, one pair per oocyst, with one or more oocysts per mosquito, and possible selfing (recombination between genetically identical parasites). Each pair generates four haploid meiotic offspring. Each offspring then replicates asexually, generating thousands of sporozoites (mosquito-to-human infectious stage) that hatch out of the oocyst/s and migrate to the salivary glands of the mosquito (Smith and Barillas-Mury, 2016). The Pv3R model is built partly around the consequent fact that a single mosquito inoculate can contain parasites that are strangers, clones and siblings, including full siblings from the same zygote (meiotic siblings) and half siblings (Figure A1, which is an adaptation of Figure 2 of (Wong et al., 2018)).

### Recurrence

The Pv3R model is articulated around three processes that each generate recurrent blood-stage *P. vivax* parasites: reinfection generates blood-stage parasites derived from a new inoculation, relapse generates blood-stage parasites derived from one or more past inoculations, recrudescence generates parasites derived from the blood-stage parasites in the directly preceding infection (Figure 1). Each inoculation contains one or more parasite broods (a collection of parasites produced when one or more oocysts hatch collectively in a mosquito). Each brood contains a collection of clones, strangers, and siblings, which include meiotic siblings from the same zygote and half siblings, as stated above. As such, the 3Rs can be reframed in terms of generating parasites that are clones, strangers and siblings of parasites in preceding blood-stage infection/s: reinfections generate strangers of parasites in preceding infections; relapses generate collections of clones, siblings and strangers of parasites in preceding infections; recrudescences generate clones of the parasites in the directly preceding infection.

**Figure 1:**
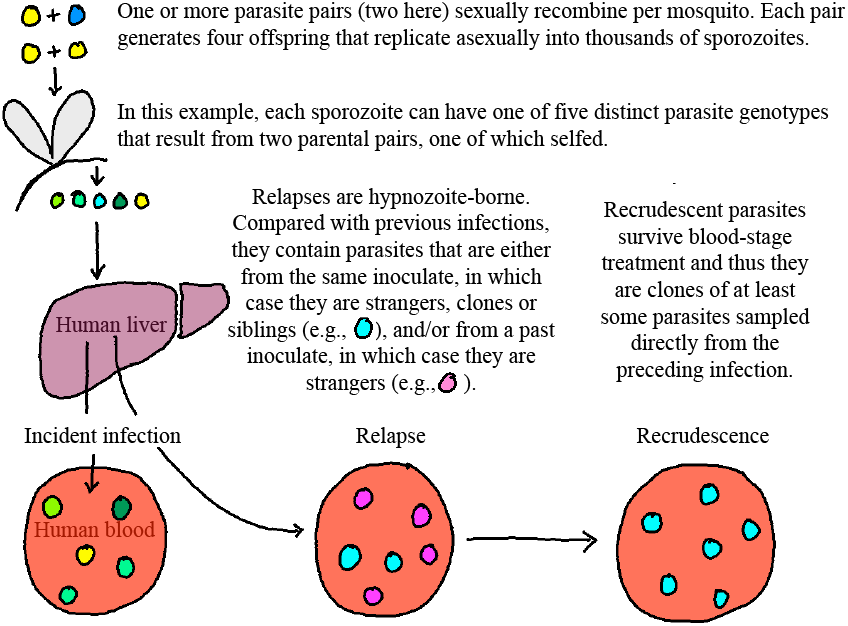
Epidemiology of *P. vivax* recurrence.

## 3 The model

### 3.1 Simplifying reality

Under the model we assume clones have relatedness one, i.e. we ignore *de novo* mutations and ectopic recombination between *var* genes etc. We assume both strangers and the parental genotypes of siblings are unrelated. This is a departure from the model in (Taylor et al., 2019), which features a background relatedness fudge factor *α* ∈ [0, 0.5]. In the Pv3R model, we considered including the inbreeding coefficient *f* ∈ [0, 1], which has an longstanding definition, that of a correlation between gametes (Hill, 1996).^1^ However, we decided against including *f* for the following three reasons. First, the inclusion of *f* severely complicates the computation of the probability of an identity-by-descent partition given a relationship graph. Second, work by Somya Mehra has shown that background relatedness is encoded in sample allele frequencies, which we are bound to use. As such, if we were to include *f*, we would be liable to overcompensate for background relatedness. Third, even if we were able to include *f*, its singular value is inadequate in that it cannot capture the fact that malaria parasites both brood and non-brood mate, i.e., they are not panmitic. We ignore meitotic siblings and half siblings, assuming all siblings have relatedness 0.5, which implies all parental parasites are unrelated (as stated above).

Having redefined the 3Rs in terms of processes that generate clones, strangers and siblings, we compute the probability of a recurrent state (recrudescence, relapse or reinfection) conditional on all data for a given study participant by summing over latent graphs of clonal, sibling and stranger relationships between genotypes within and across infections. We assume a uniform prior distribution over graphs compatible with a given recurrent state: all graphs are compatible with relapses; all graphs with clonal edges connecting genotypes in the specified recurrence to genotypes in the directly preceding infection are compatible with recrudescence; all graphs with only stranger edges connecting genotypes in the specified recurrence to genotypes in all preceding infections are compatible with reinfection. All graphs compatible with the data are summed over. This severely limits the scalability of the model: at present, it is not feasible to enumerate and sum over all graphs compatible with 11 or more genotypes in total within and across episodes. To make Pv3R scale to more markers, exact summation over graphs will be exchanged for numerical integration using, e.g., a bespoke MCMC sampler. Scalability is beyond the scope of this short report, however.

We model observed allelic data assuming perfect detection, no mutation and no error. Since no attempt is made to model genotyping error, the model is agnostic to marker type. In this document we assume markers are SNPs with alleles A, T, C and G. Although we don’t model error between observed and true alleles, we do make a distinction between observed but unphased allelic data and phased but unobserved haploid sequences of alleles (genotypes). We aim to modify the model to allowing for errors and undetected genotypes are problematic because they obfuscate clonal parasites, which include all recrudescent parasites (Figure 1), rendering recrudescence inference frail under the proof-of-principle model. The full set of model assumptions is listed below.

- No genotyping errors
- No *de novo* mutations
- No genotypes go undetected
- Markers are independent and neutral
- All siblings are regular full siblings (i.e., meitotic and half siblings are ignorable)
- Sibling parental parasites are strangers (implies panmixia, ignores brood and non-brood mating)
- Stranger parasites are unrelated (infinitely large, outbred, panmictic population)
- Mutually exclusive recurrence states
- The number of genotypes per infection is equal to the maximum number of distinct alleles seen at any marker within that infection (i.e. no undetected genotypes and sufficient diversity at at least one marker to capture all genotypes present - liable to break down for small numbers of markers)
- Missing data are ignorable

### 3.2 Mathematical description

Let *k* denote some number of recurrent vivax malaria episodes experienced by an individual. Suppose ***y*** = (***y***_*tj*_) is a matrix of *P. vivax* genetic data. Specifically, a matrix of sets, where each set, ***y***_*tj*_, contains the alleles observed in the *t*th infection at the *j* = 1, …, *m P. vivax* marker genotyped. Conditional on ***y***, Pv3R computes the posterior probability that the state of the *t*th recurrence, *s*_*t*_, is a relapse, *L*, reinfection, *I*, or recrudescence *C*. It does so by summing over all sequences of recurrent states, ***s*** = (*s*_1_, …, *s*_*k*_), in the sequence space **𝒮** where *s*_*t*_ = *L, I* or *C*, respectively,

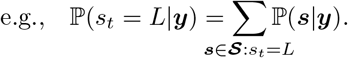

The posterior probability, ℙ(***s***|***y***), is computed assuming prior independence between recurrent states, possibly setting ℙ(*s*_*t*_) equal to the output of a time-to-event model,

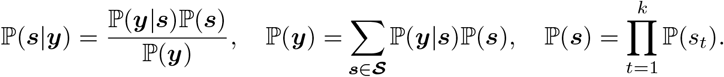

The likelihood, ℙ(***y***|***s***), sums over various latent variables. First, it sums over every matrix of phased alleles, ***a*** = (*a*_*ij*_), in matrix space **𝒜** (equation (1)); each ***a*** ∈ **𝒜** represents one way to phase the observed alleles into *i* = 1, …, *n* sequences of *j* = 1, …, *m* alleles, each sequence, ***a***_*i*·_ = (*a*_*i*1_, …, *a*_*im*_), corresponding to a genotype. Second, it sums over every genotype relationship graph, ***g***, in graph space **𝒢** (equation (3)). Third, for *j* = 1, …, *m*, it sums over every genotype identity-by-descent (IBD) partition, ***p***, in partition space **𝒫** (equation (6)). It is assumed that markers are independent conditional on a given relationship graph (equation (5)), and that the cells of an IBD partition, ***c*** ⊂ ***p***, are independent (equation (8)):

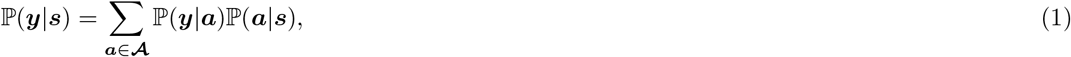

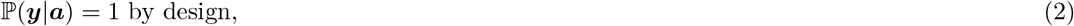

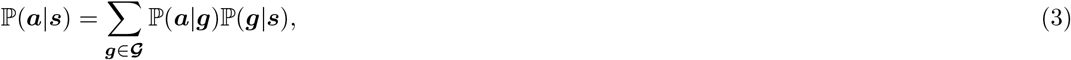

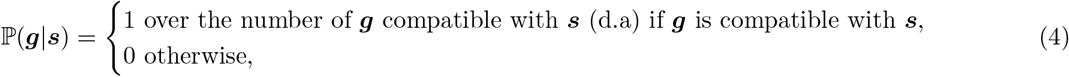

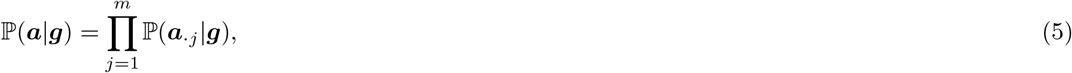

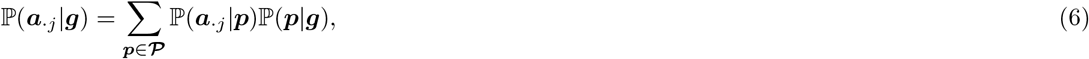

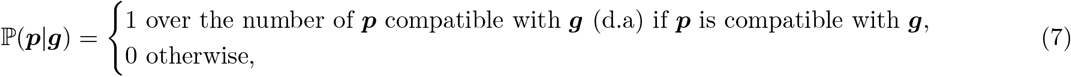

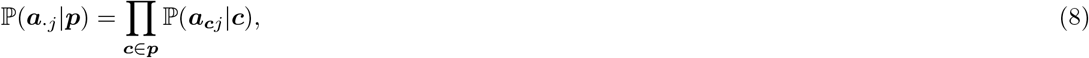

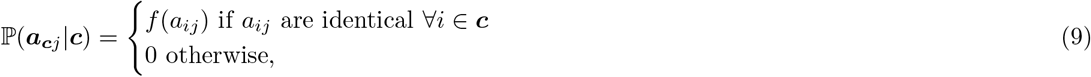

where *f* (*a*_*ij*_) is the frequency of the allele of the *i*th genotype at the *j*th marker in the matrix of phased alleles ***a***. At present, *n* is the smallest value compatible with the data assuming no genotyping errors, i.e. 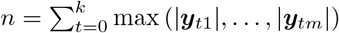, where |***y***_*tj*_| denotes the size of the set of alleles observed in the *t*th infection at the *j*th marker for *j* = 1, …, *m*. In a future version of the model, I aim to treat *n* a realised random variable. The Pv3R model is described graphically in Figure 2. A summary of all notation is provided in Table 1.

**Figure 2:**
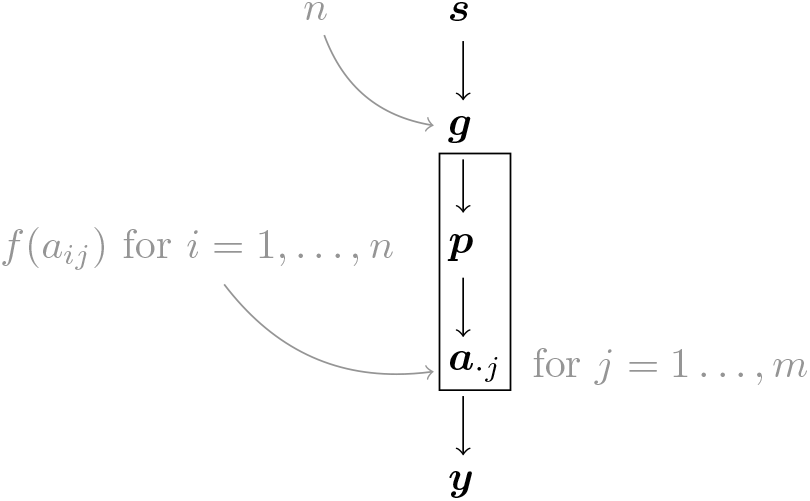
Pv3R model. Realisations of random variables in black; non-random, plug-in variables in gray.

**Figure 3:**
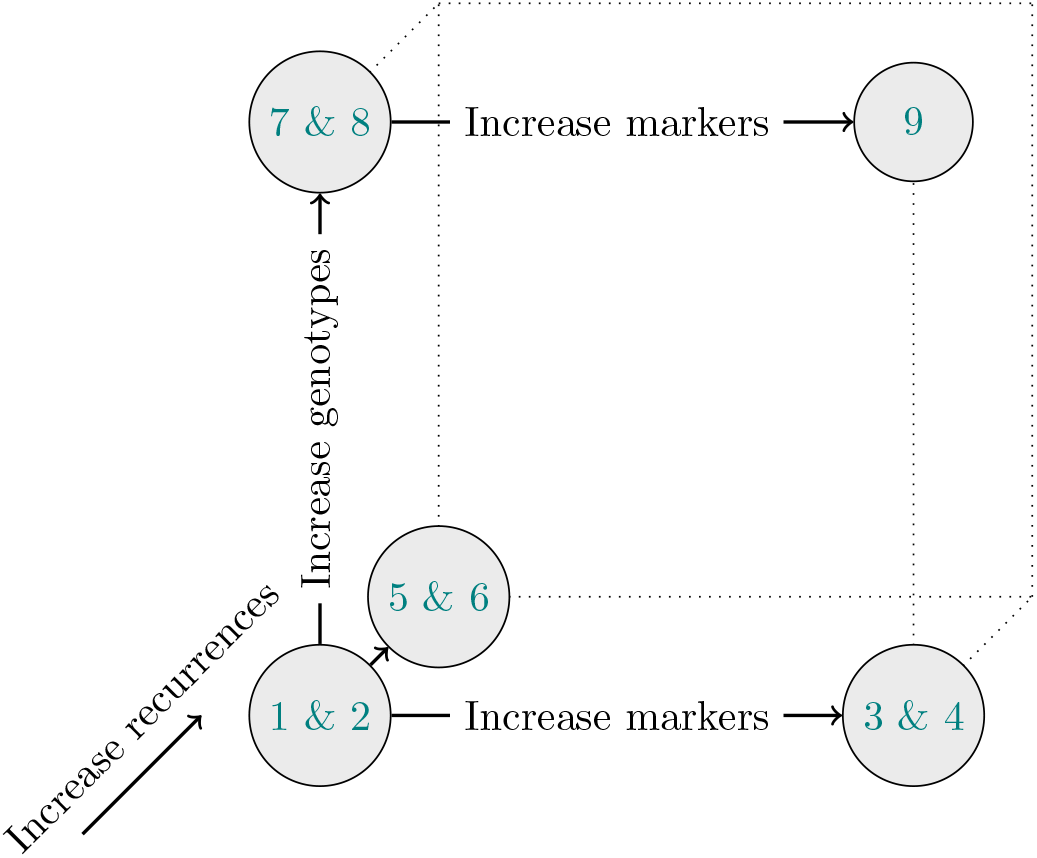
Visual summary of the examples. We start in the simplest setting (examples 4.1 & 4.2). We then add complexity in three separate directions: by increasing the number of markers (examples 4.3 & 4.4), by increasing the number of recurrences (example 4.5 & 4.6), and by increasing the number of genotypes per infection (examples 4.7 & 4.8), where the latter focuses on the computation of the probability of phased alleles given an IBD partition, to better illustrate how cells are multiplied over. Our final example addresses the need to phase, which can occur when there are multiple genotypes per infection and multiple markers typed (example 4.9).

**Table 1:** Table of notation. Bold used to distinguish vectors, matrices, sets and subsets from scalars. In the examples, roman numerals are used to enumerate members of discrete spaces (***a*** ∈ **𝒜, *p*** ∈ **𝒫, *g*** ∈ **𝒢**) over which we sum. Arabic numerals are used to index rows and columns etc.

|  |  |
| --- | --- |
| $k$ | Number of recurrent vivax malaria episodes. |
| $m$ | Number of markers genotyped. |
| $n$ | Number of parasite genotypes within and across episodes. |
| $t = 0, \dots, k$ | Index over episodes: $t = 0$ is the incident episode and $t > 0$ are recurrences. |
| $i = 1, \dots, n$ | Index over parasite genotypes. |
| $j = 1, \dots, m$ | Index over markers genotyped. |
| $\mathbf{s} = (s_1, \dots, s_k)$ | A 1-by-$k$ vector of recurrent states in sequence space $\mathcal{S}$, where $s_t \in \{L, C, I\}$ for $t = 1, \dots, k$ recurrences and where $L$ is a relapse, $C$ is a recrudescence, and $I$ is a reinfection. |
| $\mathbf{y} = (\mathbf{y}_{tj})$ | A $(1+k)$-by-$m$ matrix of sets of alleles observed in the $t$th infection at the $j$th marker. |
| $\mathbf{a} \in \mathcal{A}$ | A $n$-by-$m$ matrix of phased alleles in matrix space $\mathcal{A}$. |
| $\mathbf{g} \in \mathcal{G}$ | A graph in graph space $\mathcal{G}$, whose edges are relationships between parasite genotypes (vertices), with either sibling or stranger relationships between parasite genotypes within infections, and clonal, sibling or stranger relationships between parasite genotypes across infections; $\mathbf{g}$ can also be expressed as a $n$-by-$n$ matrix. |
| $\mathbf{p} \in \mathcal{P}$ | A partition in partition space, $\mathcal{P}$, partitioning $n$ genotypes into IBD clusters; $\mathbf{p}$ can also be expressed as a cluster graph or as an $n$-by-$n$ matrix. |
| $\mathbf{c} \in \mathbf{p}$ | An IBD cluster within an IBD partition (a cell in the partition $\mathbf{p}$). |
| $f(a_{ij})$ | Frequency of the allele of the $i$th genotype at the $j$th marker in $\mathbf{a}$. |

### 3.3 How the current model improves upon the previous one

The Pv3R model improves upon the model published in 2019 (Taylor et al., 2019), which over counts frequencies disproportionately, due to an independence assumption. For example, in the previous version of the model, if a person has three infections (one incident, two recurrent) in each of which the same allele, *A* say, is detected at a single marker genotyped, then for *α* = 0, the likelihood of the data given twice reinfection and twice recrudescence is *f* (*A*)^6^ and *f* (*A*)^3^, respectively, where *f* (*A*) is the frequency of the allele *A*. However, if reinfection is considered equivalent to drawing independently at random from the population, and if recrudescence is considered cloning with probability one, the likelihood given twice reinfection and given twice recrudescence should be *f* (*A*)^3^ and *f* (*A*), respectively, as it is in the Pv3R model (example 4.6). To prevent over counting, instead of taking a product over edges of relationship graphs (equation 8 of the supplement of (Taylor et al., 2019), which amounts to assuming independent edges), the Pv3R model takes a product over cells of IBD partitions (equation (8)). For each cell, we count one allele frequency if all alleles belonging to the genotypes within the cell at the *j*th marker are the same, and zero otherwise (equation (9)).

### 3.4 Decomposing the phasing space

Because relationships are genotype-level attributes, in the description above and in one illustrative example below, we consider it more intutitive to first phase the data into genotypes and then sum over relationship graphs between genotypes. Consequently, the likelihood involves enumerating over the matrix space **𝒜** (equation (1)), whose cardinality grows exponentially with the number of markers. In practice, phasing does not limit the scalability of the model, however, because, assuming markers are independent given the relationship graph, we can decompose the phasing space such that the enumeration over allele assignments is linear in the number of markers. Equations (1)–(4) can be expressed as

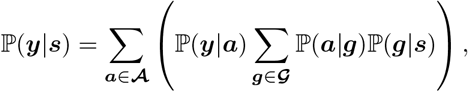

which we rearrange into

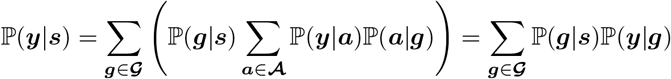

by swapping the summations, where

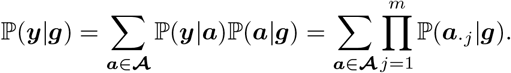

In the last equality, we use the assumption of marker independence (equation (5)), and omit the term ℙ(***y***|***a***) as it is equal to 1 by design (equation (2)).

We now focus on how to rewrite ℙ(***y***|***g***) in a way that avoids explicitly enumerating over **𝒜**. For each *j* = 1, …, *m*, let **𝒜**_*j*_ be the set of possible values of ***a***_·*j*_ (allele assignments for marker *j*) consistent with the observed data **y**. The key observation is that **𝒜** decomposes into a Cartesian product over **𝒜**_1_, …, **𝒜**_*m*_. This implies that

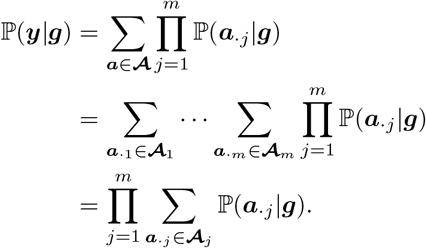

Consider an example where two alleles are observed at two of three markers genotyped in an incident infection, and the same scenario repeats for a recurrent infection, where one of the two markers are different:

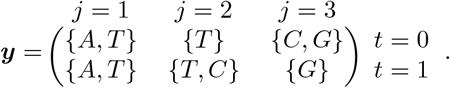

We assume the most parsimonious explanation of the data: the number of genotypes in the first and second infections are both two. There are four ways to phase allelic data into *n* = 4 genotypes:

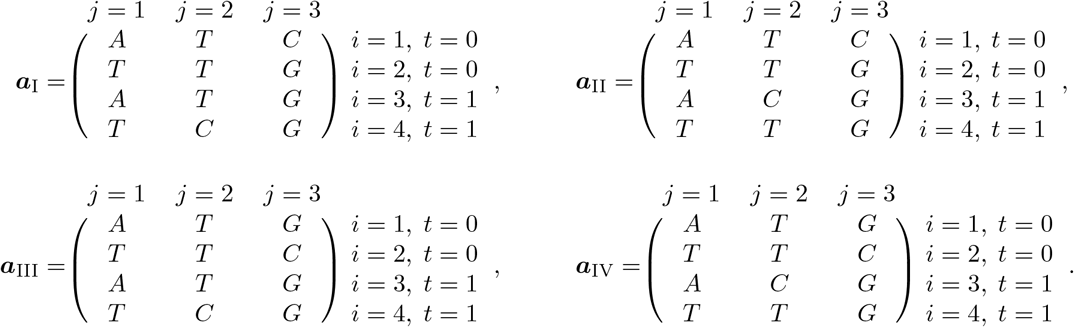

Let 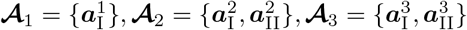, where

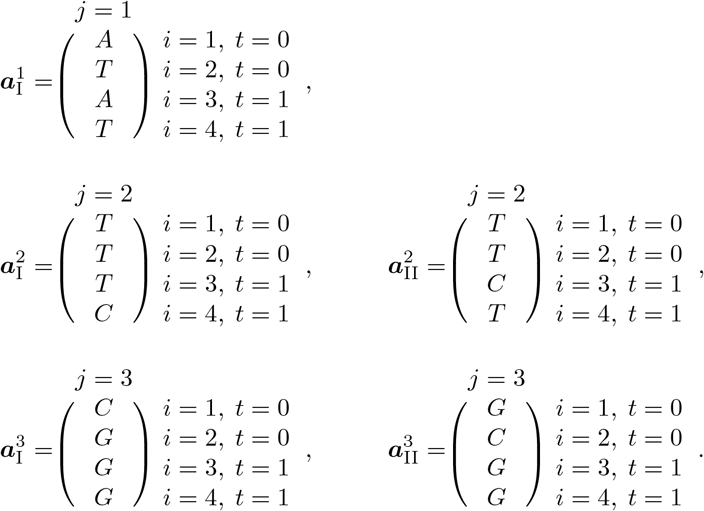

In the original formulation, we would have

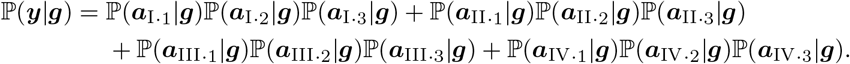

We now substitute

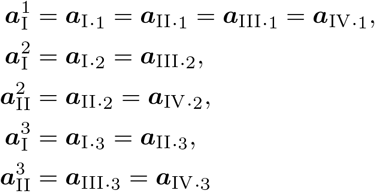

and arrive at the factorisation

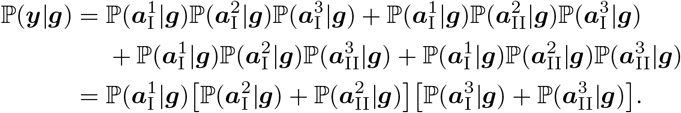

## 4 Examples

### 4.1 Single heterologous marker

#### Observed and phased alleles

Suppose a single allele is observed at a single marker, *m* = 1, indexed by *j*, genotyped in an incident infection, *t* = 0, and single recurrent infection, *t* = 1. Since we detect only one allele per infection we assume there is only one genotype per infection, indexed by *i*, and only one way to phase the observed alleles, e.g.,

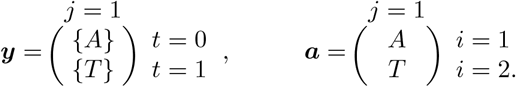

#### Relationship graphs

To compute the posterior probability of the single recurrent state, *s*, being either a relapse, *L*, reinfection, *I*, or recrudescence *C*, we sum over three graphs (***g***_I_, ***g***_II_, ***g***_III_) of relationships between parasite genotypes, each genotype 1 and 2,

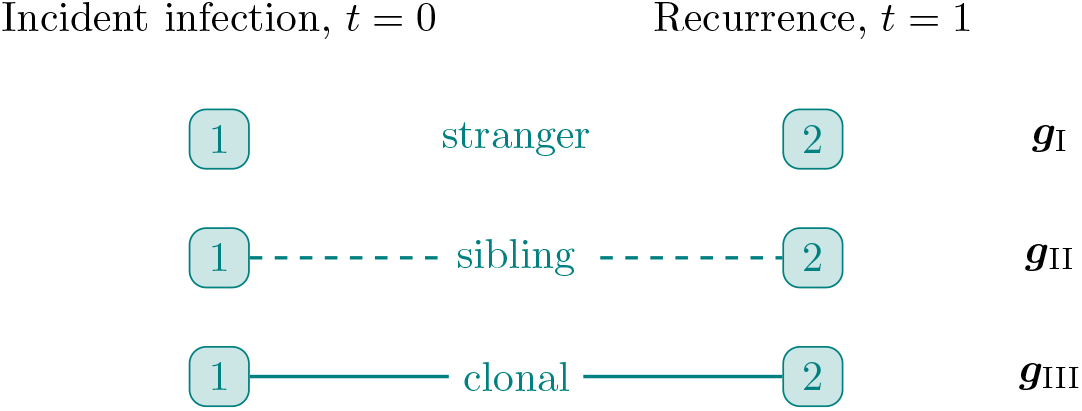

#### IBD partitions

For each relationship graph at each marker, which in the case of *m* = 1 is a single marker, we sum over two IBD partitions of genotypes 1 an2, ***p***_I_ = {{1}, {2}} and ***p***_II_ = {{1, 2}} that have two and one cell, respectively, and correspond to the following two IBD graphs.

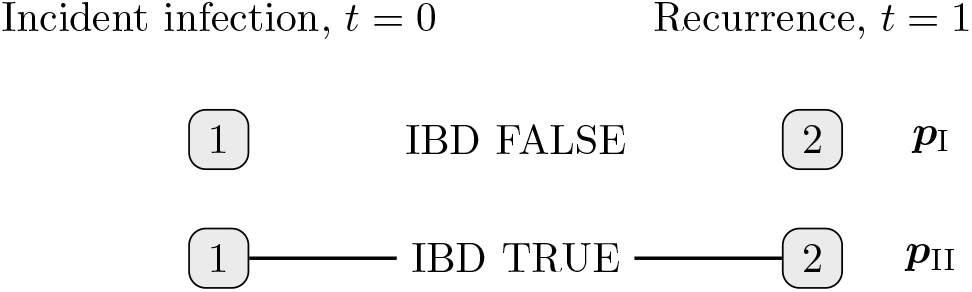

#### Likelihood

As there is one way to phase the observed alleles and *m* = 1, ℙ(***y***|*s*) = ℙ(***a***|*s*) = ℙ(***a***_·1_|*s*) where

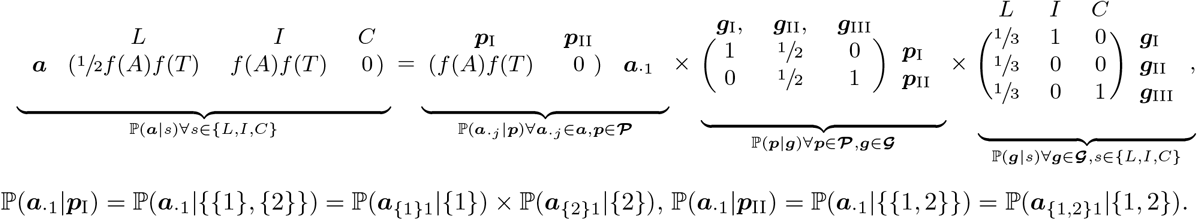

#### Posterior

For a uniform prior on *s*, i.e., ℙ(*s*) = ⅓∀*s* ∈ (*L, I, C*), ℙ(*y*) = ½*f* (*A*)*f* (*T*), such that ℙ(*s* = *L*°***y***) = ⅓, ℙ(*s* = *I*°***y***) = ⅔ and ℙ(*s* = *C*|***y***) = 0.

### 4.2 Single homologous marker

For ***y*** = ({*A*}, {*A*})^*T*^ and a uniform prior on s, ℙ(*y*) = ½*f* (*A*)^2^ + ½*f* (*A*), such that ℙ(*s* = *L*|***y***) = ⅓, ℙ(*s* = *I*|***y***) = ⅔*f* (*A*)(*f* (*A*) + 1)^−1^ and ℙ(*s* = *C*|***y***) = ⅔(*f* (*A*) + 1)^−1^ following

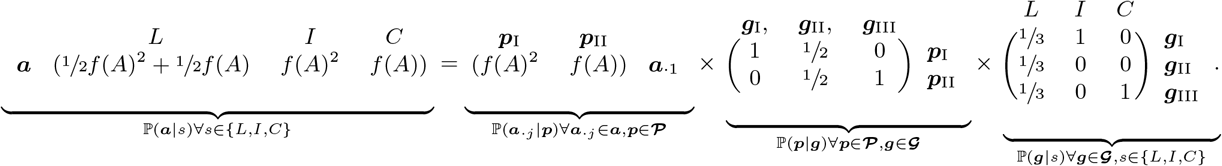

### 4.3 Multiple partially heterologous markers

#### Observed and phased alleles

In the simplest example a single allele is observed per marker genotyped in an incident and single recurrent infection. Since we detect only one allele per infection we assume there is only one genotype per infection and thus there is only one to phase the observed alleles, e.g.,

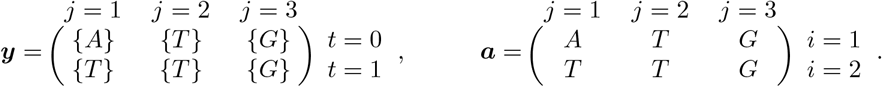

#### Relationship graphs and IBD partitions

We sum over the same three graphs of relationships between genotypes and the same two IBD partitions as in example 4.1.

#### Likelihood

The likelihood computation is similar to examples 4.1 & 4.2, but because there are multiple markers, which are conditionally independent given the relationship graphs, a product over markers is taken before summing over relationship graphs. In addition, alleles are indexed by *j*, since a given allele at one locus might have a different frequency at another locus.

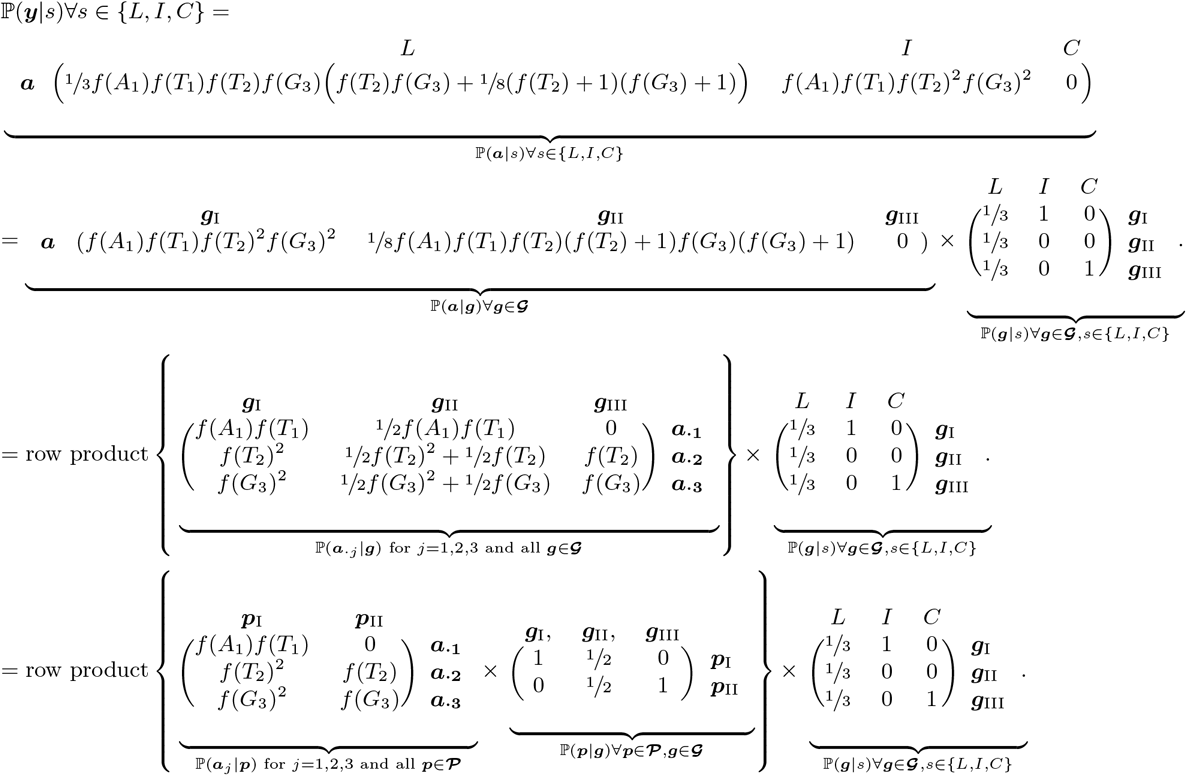

#### Posterior

For a uniform prior on s,

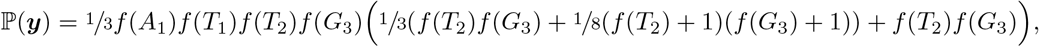

such that

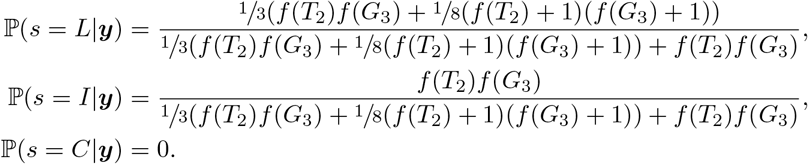

### 4.4 Multiple exclusively homologous markers

When markers are homologous over infections, ℙ(*s* = *C*|***y***) *>* 0. For example, if

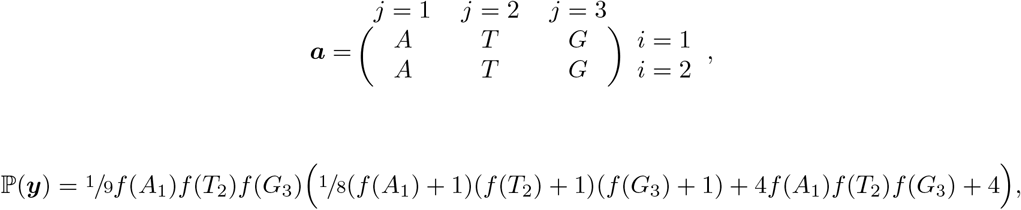

such that

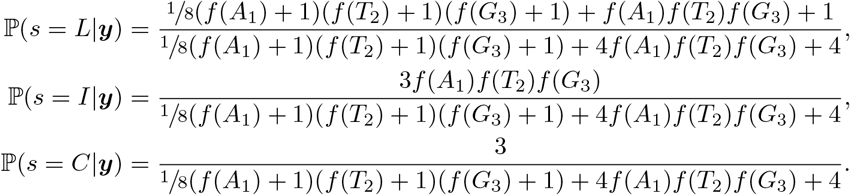

because

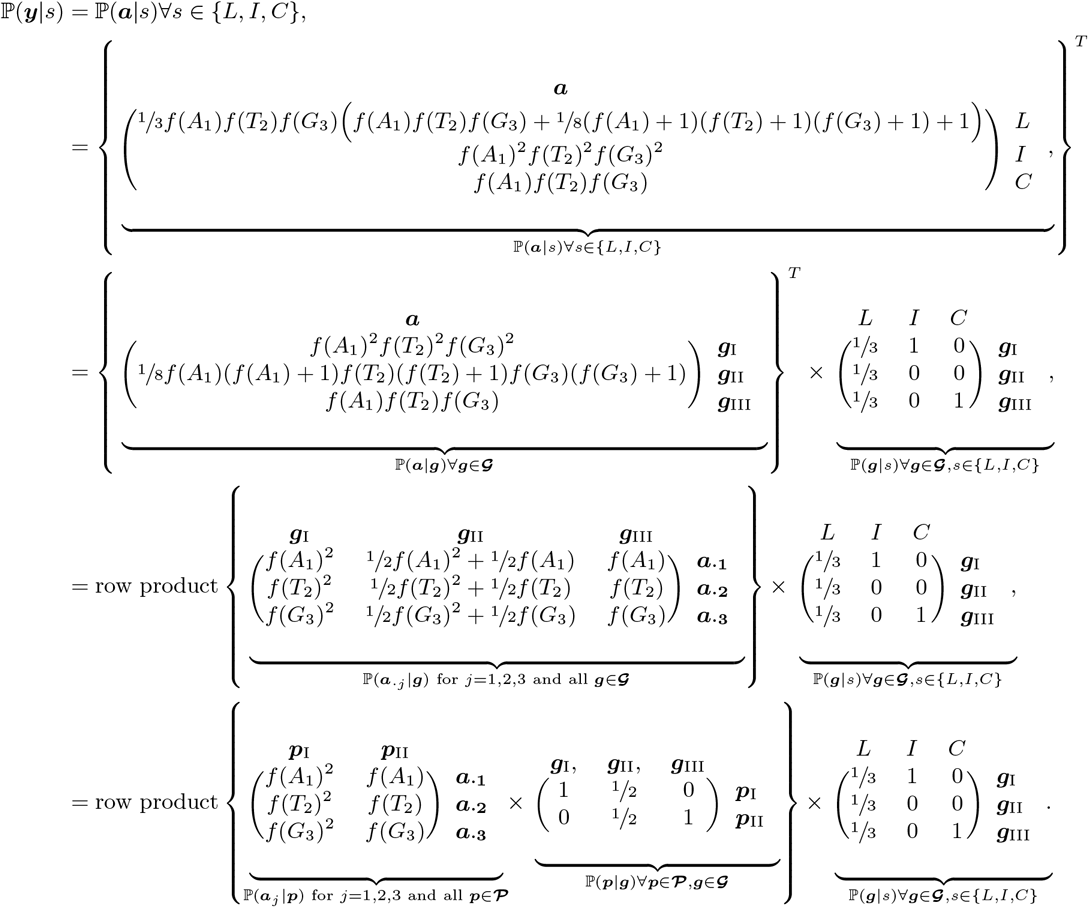

### 4.5 More than one recurrence: partially heterologous markers

#### Observed and phased alleles

In the simplest example a single allele is observed at one marker genotyped in an incident infection *t* = 0 and in *t* = 1, 2 recurrent infections. Since we detect only one allele per infection we assume there is only one genotype per infection, indexed by *i*, and thus there is only one way to phase the observed alleles, e.g.,

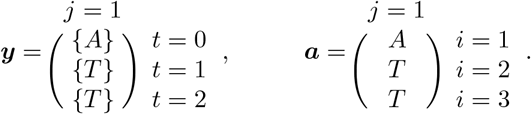

#### Relationship graphs and IBD partitions

For the simplest example with two recurrences, we sum over 12 relationship graphs, shown below. For each relationship graph, at the single *j* = 1 marker, we sum over five IBD partitions, shown directly below, which can also be depicted as IBD graphs as follows.

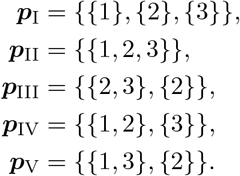

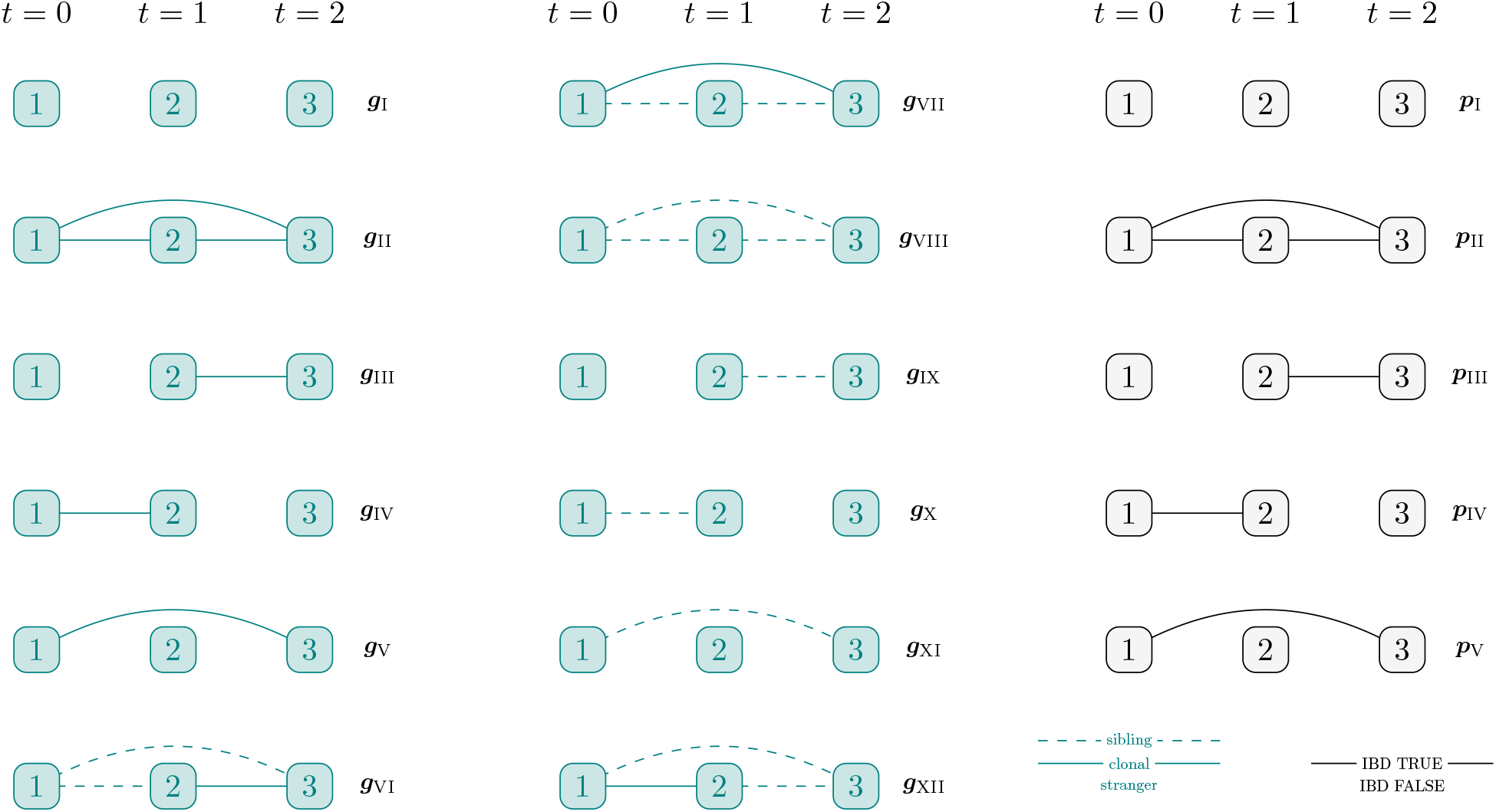

#### Likelihood

Remembering that in linear algebra (*CBA*)^*T*^ = (*A*^*T*^ *B*^*T*^ *C*^*T*^),

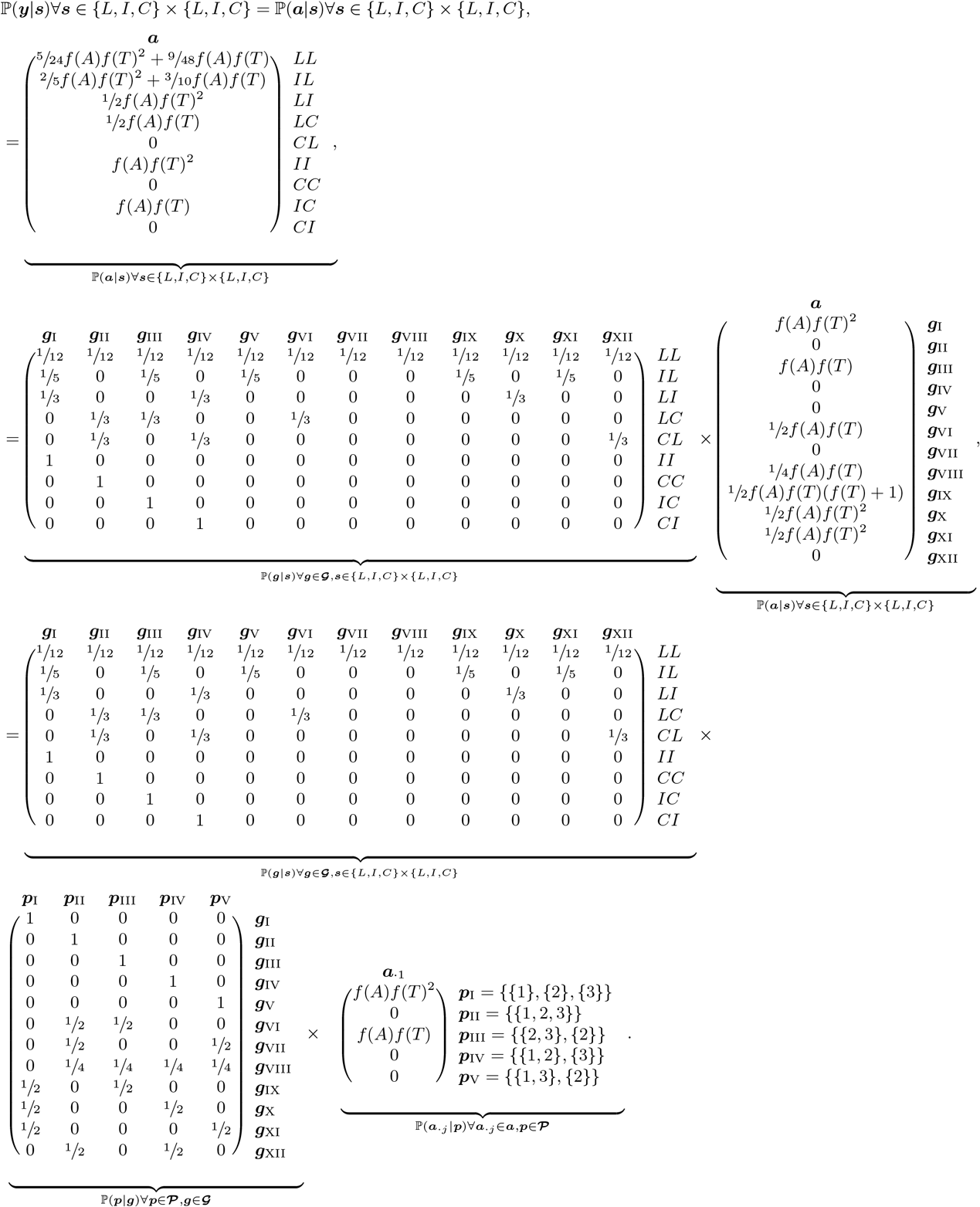

#### Posterior

For a uniform prior on 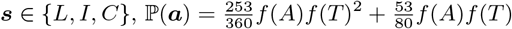 and

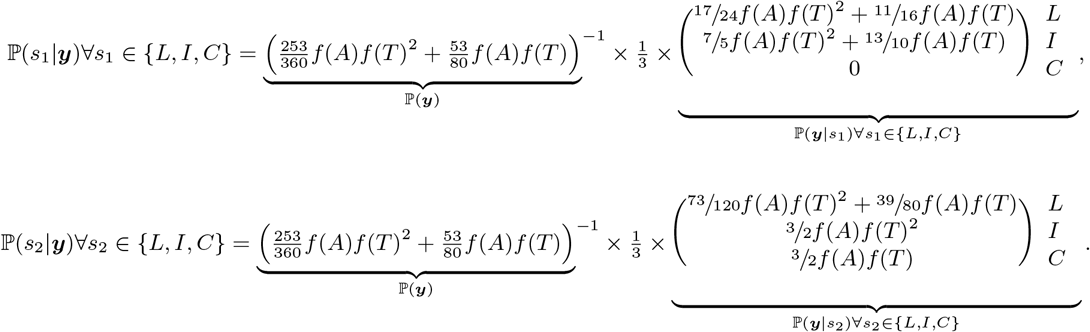

### 4.6 More than one recurrence: exclusively homologous markers

If instead ***y*** = ({*A*}, {*A*}, {*A*})^*T*^, ℙ(***y***|***s***) = ℙ(***a***|***s***), which for ***s*** = *II* and ***s*** = *CC*

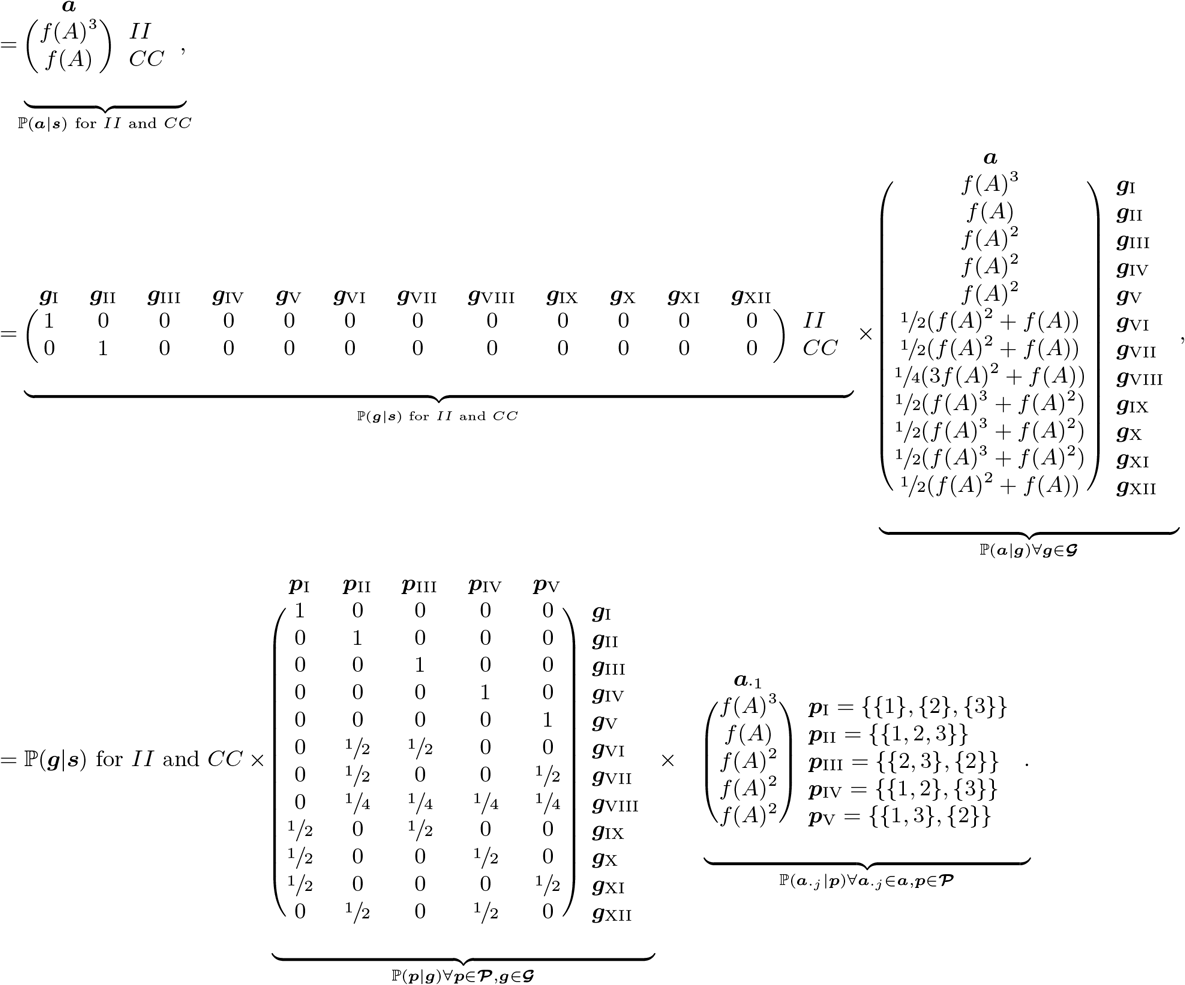

### 4.7 Simple example for more than one genotype per infection

#### Observed and phased alleles

In a simple example two and one alleles are observed at a single marker genotyped in an incident and single recurrent infection, respectively. We assume the most parsimonious explanation of the data: the number of genotypes in the first and second infections are two and one, respectively. There is no-longer a one-to-one mapping between genotypes and infections over time. However, there remains one matrix of phased alleles compatible with the observed alleles, e.g.,

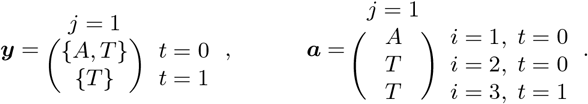

#### Relationship graphs and IBD partitions

There are nine relationship graphs (a subset of those in example 4.5 because clonal relationships within infections are disallowed) to sum over and five IBD partitions (the same as those in example 4.5), corresponding to the same five IBD graphs as follows.

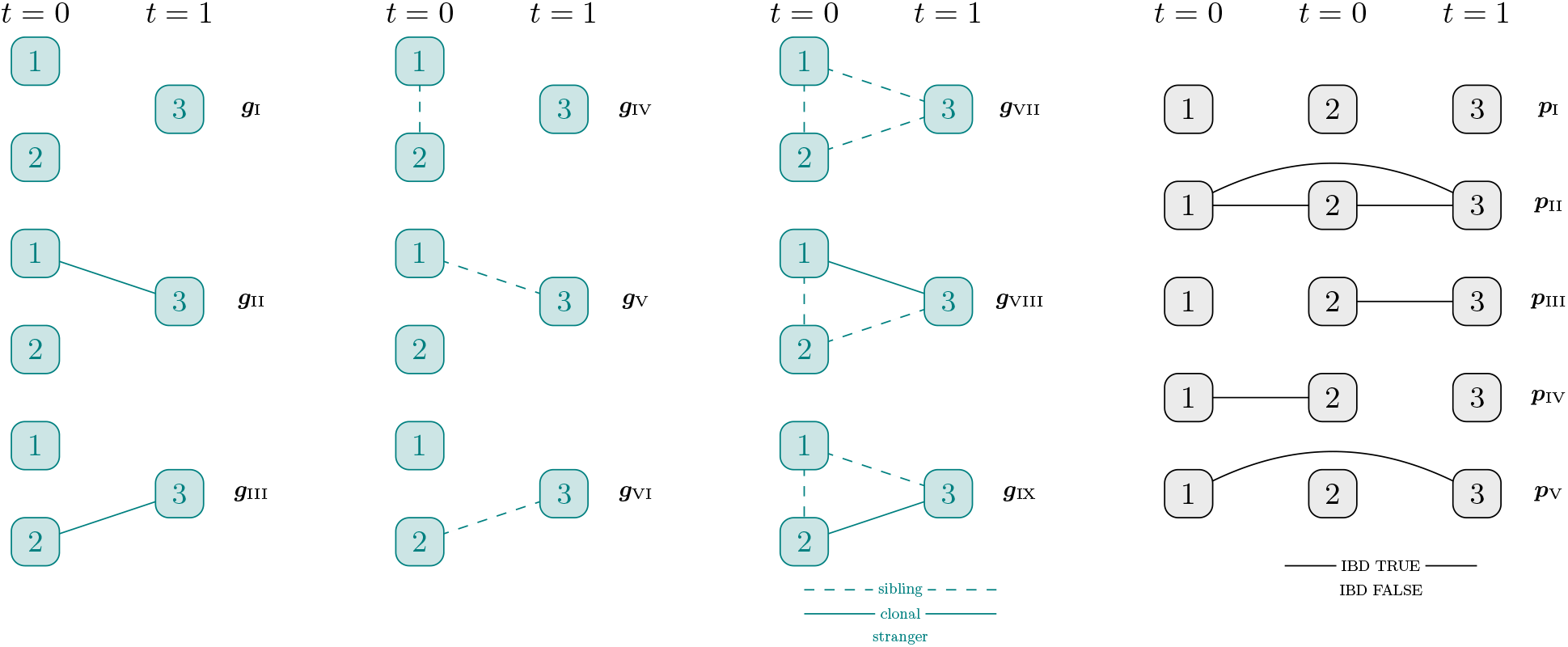

#### Likelihood

Remembering that in linear algebra (*CBA*)^*T*^ = (*A*^*T*^ *B*^*T*^ *C*^*T*^),

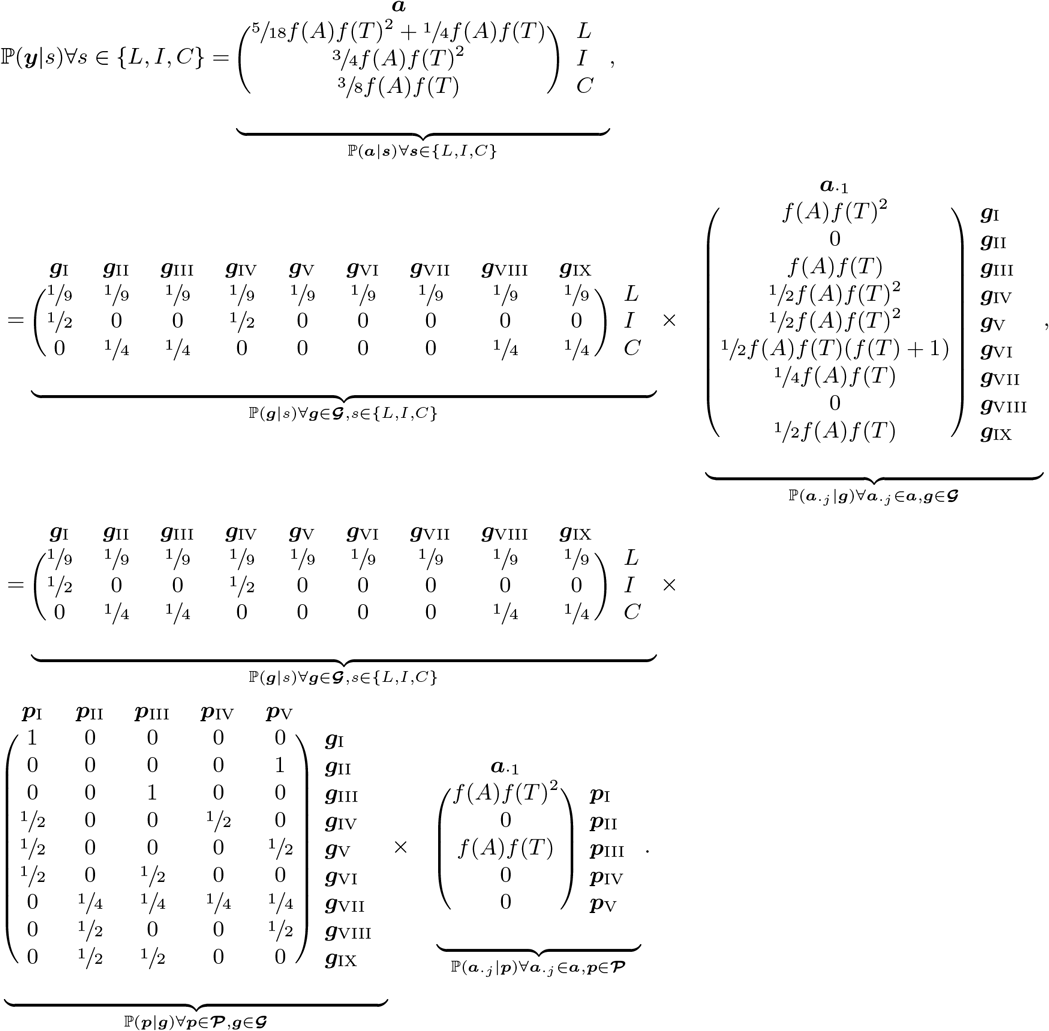

Note that if the data for the incident infection and the recurrence were reversed, e.g. if ***y***_*t*=0_ = {*T* } and ***y***_*t*=1_ = {*A, T* }, the posterior probability of recrudescence would be zero because recrudescences must have the same or fewer genotypes than the directly preceding infection.

#### Posterior

For a uniform prior on 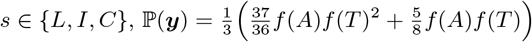 and

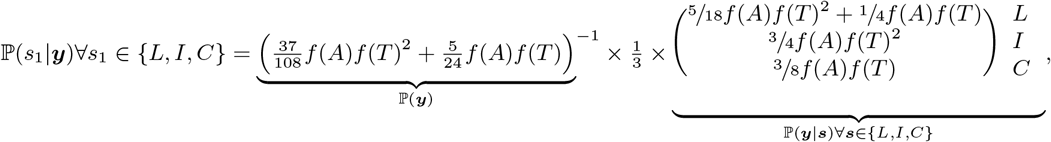

### 4.8 Simple zoomed-in example for six genotypes

#### Observed and phased alleles

Suppose the observed alleles suggest there are *n >* 3 genotypes, e.g.,

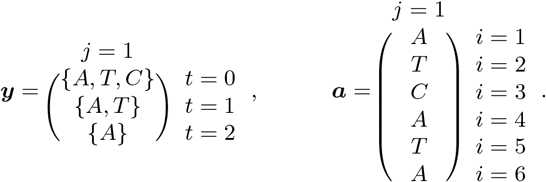

#### IBD partitions

For *n* = 6, there are 203 IBD partitions in **𝒫**. Three examples, depicted below as graphs, are:

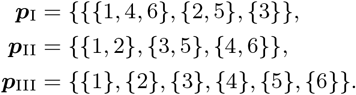

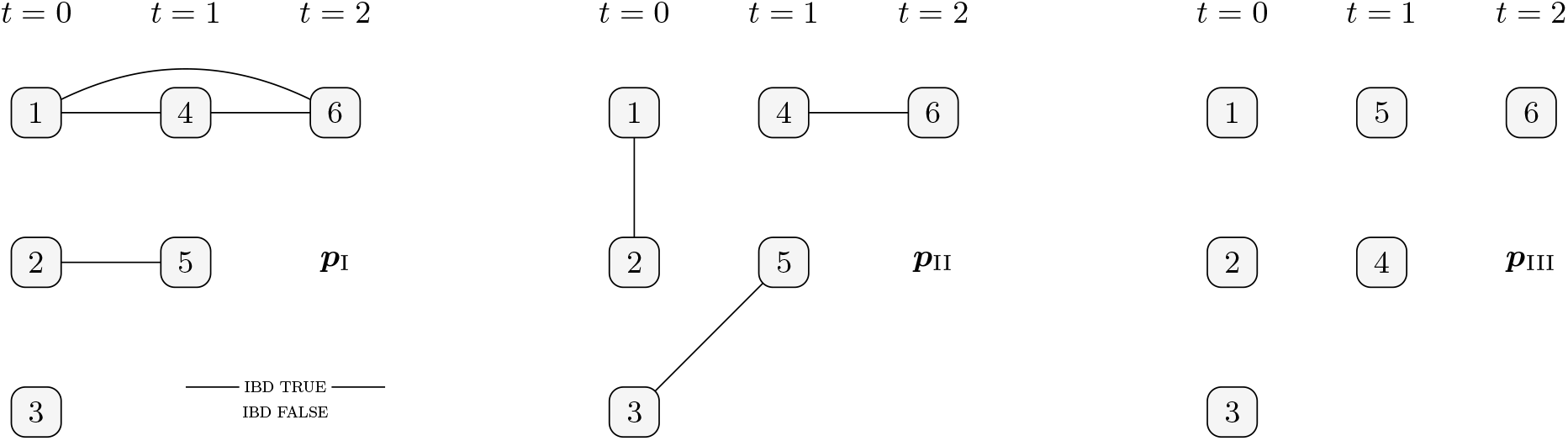

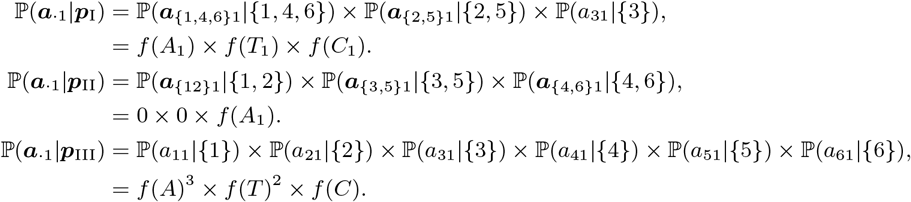

### 4.9 Simple example involving phasing

#### Observed and phased alleles

In a simple example two alleles are observed at two of three markers genotyped in an incident infection, while one allele is observed per marker genotyped in a single recurrent infection. As in example 4.7, we assume the most parsimonious explanation of the data: the number of genotypes in the first and second infections are two and one, respectively. However, there are now two ways to phase allelic data into *n* = 3 genotypes,

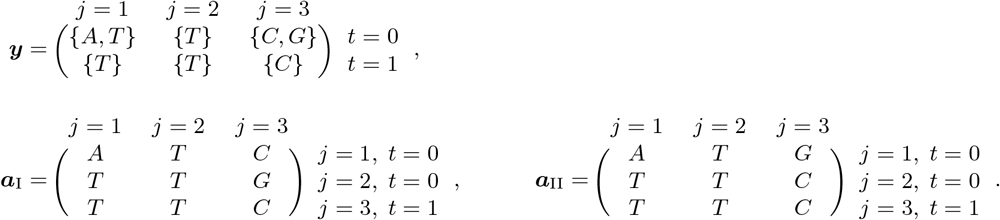

#### Relationship graphs and IBD partitions

We sum over relationship graphs and IBD partitions in example 4.7.

#### Likelihood

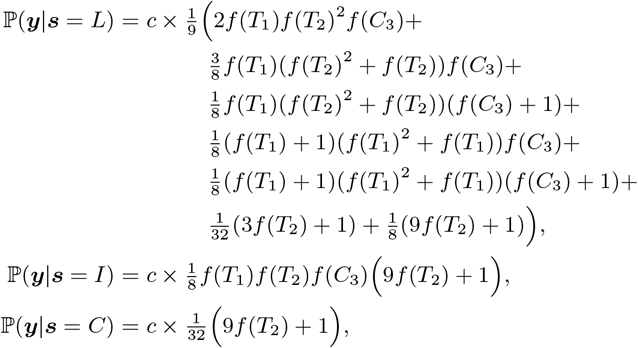

where *c* = *f* (*A*_1_)*f* (*T*_1_)*f* (*T*_2_)*f* (*C*_3_)*f* (*G*_3_).

The likelihood is a summation over ***a***_I_ and ***a***_II_:

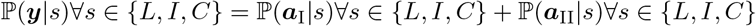

where, with differences between ***a***_I_ and ***a***_II_ highlighted in blue,

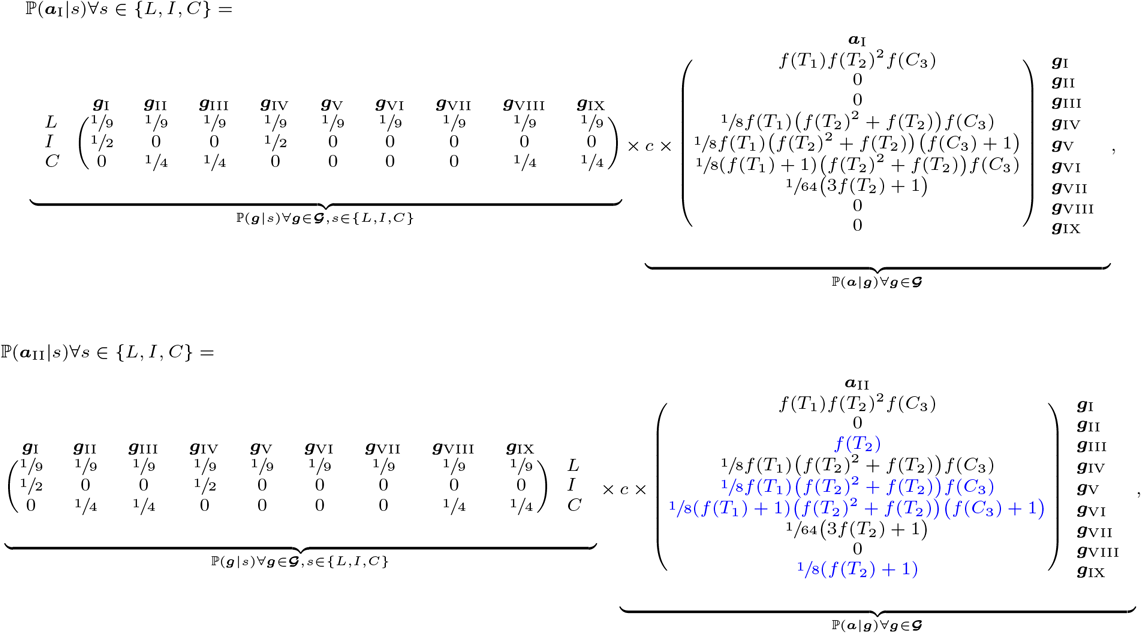

and where

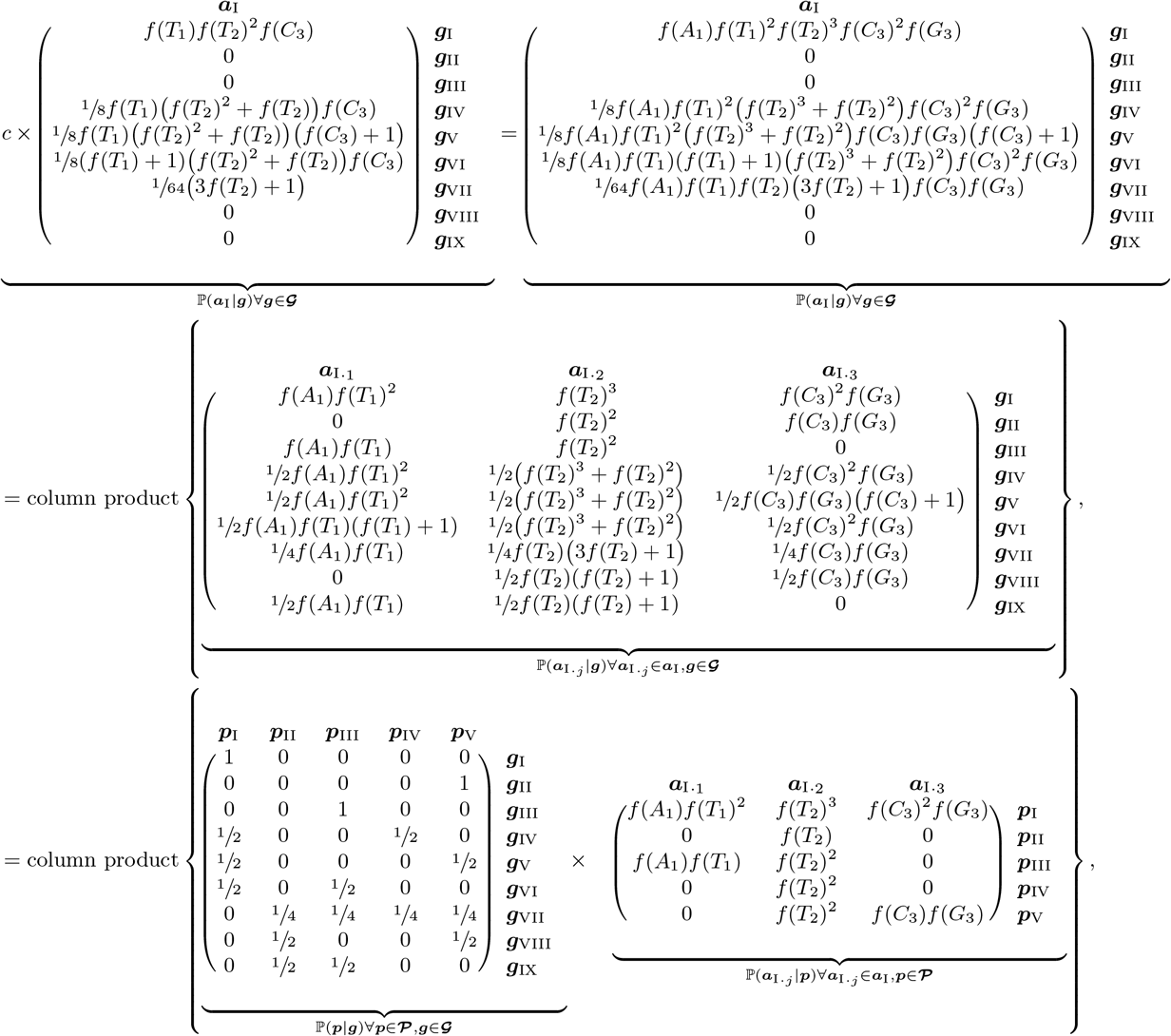

and

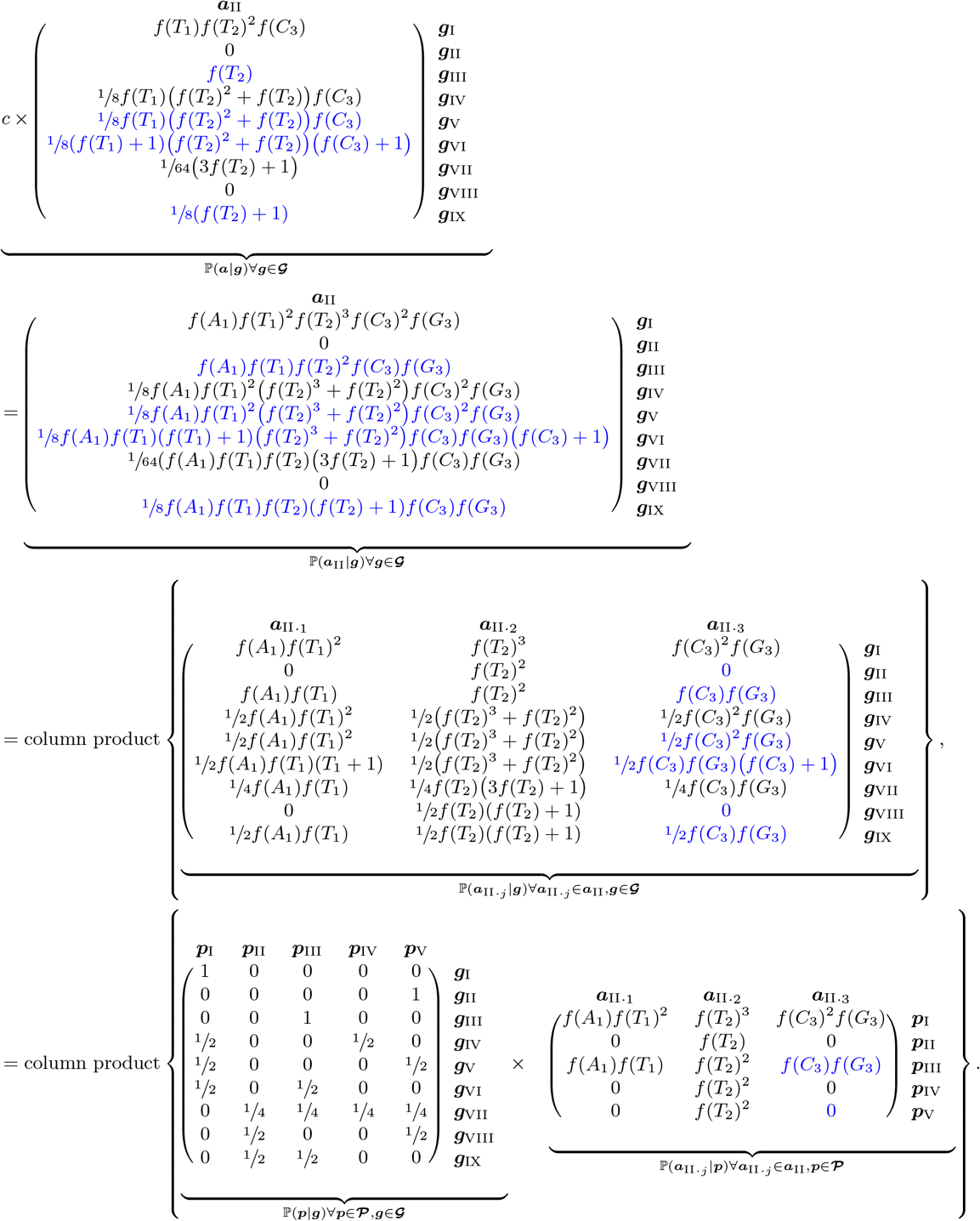

## Data Availability

All data produced in the present work are contained in the manuscript

## Appendix

**Figure A1:**
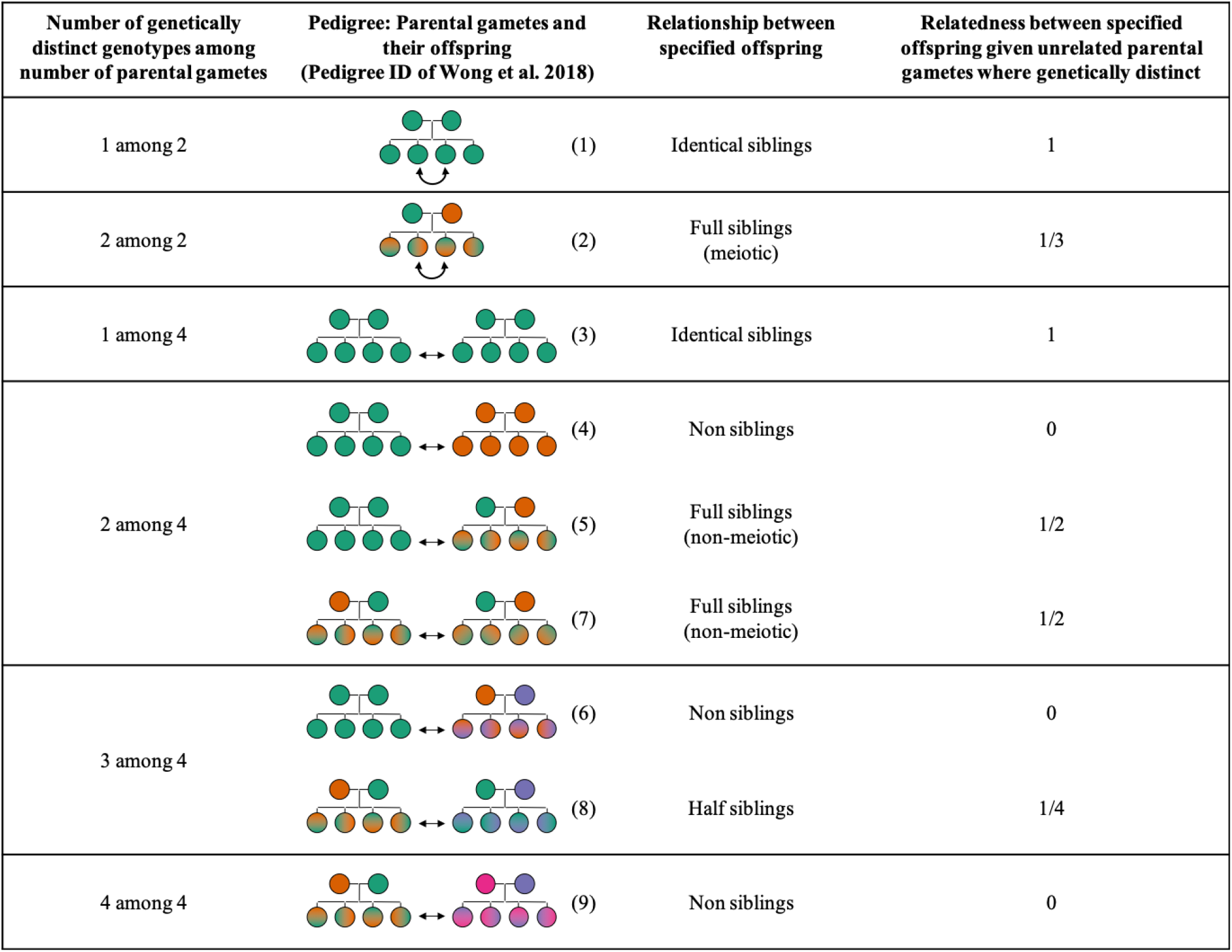
Adapted from (Wong et al., 2018). Intra-brood relationships: among the parental gametes from which a pair of sporozoites that belong to the same brood are ultimately derived, there is at most four genetically distinct genotypes (represented by different solid colours) leading to a limited set of inter-offspring (and thus inter-sporozite) relationships within a parasite brood. Note that full siblings share the same parents (represented by colour gradients) but are distinct (represented by different gradient orientations). Identical siblings and non siblings are referred to as clones and strangers in the main text. In addition to these relationships, sporozoites can be mitotic copies of one another because offspring replicate mitotically before maturing in sporozoites, and from different broods if the mosquito was superinfected by blood meals taken more than approximately an hour apart (Smith and Barillas-Mury, 2016).

## Footnotes

1 In a panmitic population with equifrequent but finite lineages, strangers have relatedness *f*, and regular full siblings with unrelated parental parasites have relatedness 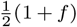, equal to twice the kinship coefficient of equation 13 in (Speed and Balding, 2015), assuming all siblings share two haploid ancestors.

